# Substance Use and Clinical Correlates of Opioid Dependence: A Multifactorial Analysis

**DOI:** 10.1101/2025.05.14.25327645

**Authors:** Hanzooma Hatwiko, David N. Masta, Salma M. Baines, Kimberley R. Kurehwatira, Prince Mulambo, Emmanuel O. Riwo, Nestorine N. Ngongo, Emmanuel Yumba, Mupuna Nature, Mundia Mwangelwa, Juliet Katongo, Natasha Chishala, Chileleko Siakabanze, Emmanuel Luwaya, Chakulya Martin, Joreen P. Povia, Sepiso K. Masenga

## Abstract

**Background:** Opioid dependence poses a growing public health challenge in low- and middle-income countries (LMICs), yet data from sub-Saharan Africa remain scarce. This study examines sociodemographic, clinical, and substance use correlates of opioid dependence among adolescents and young adults in Zambia.

**Methods:** A cross-sectional analysis of 427 medical records (aged 10–27 years) from a tertiary hospital in Southern Zambia (2022–2024) was conducted. Univariable and multivariable logistic regression models assessed associations between opioid use, sociodemographic factors, psychiatric comorbidities, and physiological markers.

**Results:** The median age was 21 years (IQR: 18–23), with 77.5% male participants. Opioid use prevalence was 15.5%, strongly associated with cannabis use (76.7% vs. 7.8%, p < 0.0001) and alcohol abuse (15.5% vs. 10.3%, p = 0.0002). Psychosis was less prevalent among opioid users (12.1% vs. 19.7%, p = 0.032). In adjusted models, systolic blood pressure (SBP) inversely correlated with opioid use (AOR = 0.80, 95% CI: 0.65–0.99; p = 0.040), while diastolic blood pressure (DBP) showed a positive association (AOR = 1.37, 95% CI: 1.03–1.81; p = 0.027). Alcohol abuse retained significance (AOR = 3.76, 95% CI: 1.47–95,874.35; p = 0.036), though wide confidence intervals indicated instability.

**Conclusion:** Opioid dependence in this Zambian cohort is closely linked to polysubstance use, particularly cannabis and alcohol. The paradoxical inverse relationship between SBP and opioid use may reflect confounding by comorbidities or antihypertensive treatment, while elevated DBP aligns with regional studies on opioid-related cardiovascular risk. Despite limitations, including sparse data and cross-sectional design, these findings underscore the need for integrated substance use and mental health interventions in LMICs. Future research should prioritize longitudinal designs and community-based sampling to address methodological gaps and inform context-specific policies.

## Introduction

Substance use disorders (SUDs), particularly opioid dependence, represent a growing global public health crisis with far-reaching medical, social, and economic consequences [1–3]. The It is estimated that over 40 million people suffer from opioid use disorders worldwide, contributing to rising overdose mortality and healthcare burdens [4]. Opioid dependence is characterized by neurobiological adaptations leading to compulsive drug-seeking behavior, tolerance, and withdrawal symptoms, often co-occurring with other psychiatric conditions and polysubstance use [5,6].

The rising prevalence of opioid misuse, particularly among adolescents and young adults, has been well-documented in high-income countries, but emerging evidence suggests increasing rates in low- and middle-income countries (LMICs) as well. This shift is particularly concerning given the limited addiction treatment infrastructure in many LMICs [7–9]. Studies indicate that early-onset substance use especially during adolescence is associated with worse long-term outcomes, including higher risks of dependence, psychiatric comorbidities, and premature mortality [10–12].

A robust body of literature demonstrates strong associations between opioid dependence and other substance use disorders, particularly alcohol and cannabis use [13]. Cannabis use disorder (CUD) has been identified as a significant risk factor for subsequent opioid misuse, possibly due to shared neurobiological pathways involving the mesolimbic dopamine system [14,15]. Additionally, individuals with opioid use disorder (OUD) frequently exhibit high rates of psychiatric comorbidities, including depression (37–54%), anxiety disorders (25–43%), and psychotic illnesses (8–15%) [16–18].

The relationship between opioid use and psychosis remains complex. While some studies suggest that opioid exposure may exacerbate psychotic symptoms, others propose that individuals with psychotic disorders may avoid opioids due to symptom exacerbation, supporting the self-medication hypothesis [19–22]. Furthermore, sociodemographic factors such as urban residence, unemployment, and lower educational attainment have been consistently linked to higher opioid misuse rates [23,24].

Despite extensive research in high-income settings, data on opioid dependence patterns in sub-Saharan Africa remain scarce. Zambia, like many LMICs, faces a growing substance abuse crisis, yet systematic studies on clinical correlates of opioid dependence are lacking. Existing research in the region has primarily focused on alcohol and cannabis use, with limited attention to opioids [25]. Furthermore, most diagnostic and treatment frameworks for OUD are derived from Western populations, raising questions about their applicability in African contexts [26,27].

This study addresses these gaps by examining the clinical and sociodemographic correlates of opioid dependence among adolescents and young adults in Zambia. Using hospital-based data, we investigate associations between opioid use, cannabis and alcohol abuse, psychiatric comorbidities, and physiological markers. Our findings contribute to the limited literature on OUD in LMICs and provide insights for regionally tailored interventions.

## Methodology

### Study Design

This study employed a cross-sectional design, performing a secondary analysis of data from a project titled “Prevalence and Factors Associated with Cannabis Use Among Adolescents and Young Adults at LUTH in Zambia.” The original study is currently under peer review (Manuscript ID: PONE-D-25-23402).

### Study Setting

The research was carried out at the Levy Mwanawasa University Teaching Hospital (LUTH), a tertiary-level referral facility catering to Zambia’s Southern Province. Data were obtained from the psychiatry department’s records, as they maintain structured documentation of substance use disorders, particularly among adolescents and young adults.

### Eligibility and Recruitment

The psychiatry department’s records contained 2,500 available case files. From these, we systematically reviewed 1,500 records of adolescents and young adults (aged 10–27 years) who received care between 2022 and 2024, as documented in the original study on cannabis use prevalence at LUTH (manuscript under review, PONE-D-25-23402). Following screening, 427 files met the inclusion criteria and were included in the final analysis. A total of 1,073 records were excluded due to incomplete data, including missing demographic variables (age, sex, residence) or unspecified diagnosis.

### Data Collection

A structured review of medical records was performed over a six-week period (March 1–April 15, 2025). To maintain data integrity, the extraction process was carried out by trained research assistants following standardized protocols.

### Study variables

The principal outcome measure in this study was opioid use. Opioid Use Disorder (OUD) was clinically characterized by a pattern of opioid consumption marked by a compelling urge or strong compulsion to use opioids, impaired control over opioid intake, continued use despite the occurrence of negative consequences, and the prioritization of opioid use over other responsibilities and activities [28].

The independent variables were classified into two main domains: sociodemographic and clinical factors. Sociodemographic variables comprised age, sex, residential setting (urban or rural), employment status, level of educational attainment, number of children, and history of criminal behavior. Clinical variables included developmental milestone achievement, degree of insight into mental illness, systolic and diastolic blood pressure (SBP and DBP), alcohol and opioid use, duration of substance use, current pharmacological interventions, and the presence of psychiatric comorbidities such as psychosis, depression, and anxiety. Additional clinical parameters comprised diagnostic history, serum creatinine concentration, alanine aminotransferase (ALT) levels, white blood cell (WBC) count, neutrophil count, lymphocyte count, monocyte count, red blood cell (RBC) count, haemoglobin concentration, platelet count, and findings from urine toxicology screening.

### Data Analysis

Data were exported from the REDCap application into Microsoft Excel 2013 for cleaning and coding. Subsequently, statistical analysis was performed using StatCrunch. Descriptive statistics were used to summarize categorical variables as frequencies and percentages, while continuous variables were described using medians and interquartile ranges. The Shapiro–Wilk test was employed to assess the normality of continuous data. Associations between categorical variables were examined using the chi-square test, and differences between two medians were assessed using the Wilcoxon rank-sum test. Univariable and multivariable logistic regression analyses were conducted to identify factors associated with cannabis use.

### Ethics approval and consent to participants

Ethical approval for this study was granted by the Mulungushi University School of Medicine and Health Sciences Research Ethics Committee (Reference Number: SMHS-MU2-2024-67) on 6 June 2024. Additionally, authorization to access patient records was obtained from the administration of LUTH. All data collected and analyzed were de-identified to maintain strict confidentiality. No personally identifiable information was extracted or recorded in the data collection tool at any stage of the study. This project utilized secondary data; hence, written or verbal informed consent was not required and was formally waived by the ethics committee.

The reporting of this study adhered to the Strengthening the Reporting of Observational Studies in Epidemiology (STROBE) guidelines, as detailed in Supplementary File S1.

## Results

Out of 2,500 files eligible for review, 1,500 medical records were examined. Among these, 427 records satisfied the inclusion criteria and were subsequently analyzed. The remaining 1,073 records were excluded owing to insufficient data, including missing demographic details (e.g., age, sex, residence) or incomplete clinical information (e.g., diagnosis or outcome), as illustrated in **Figure 1**.

**Figure 1.**
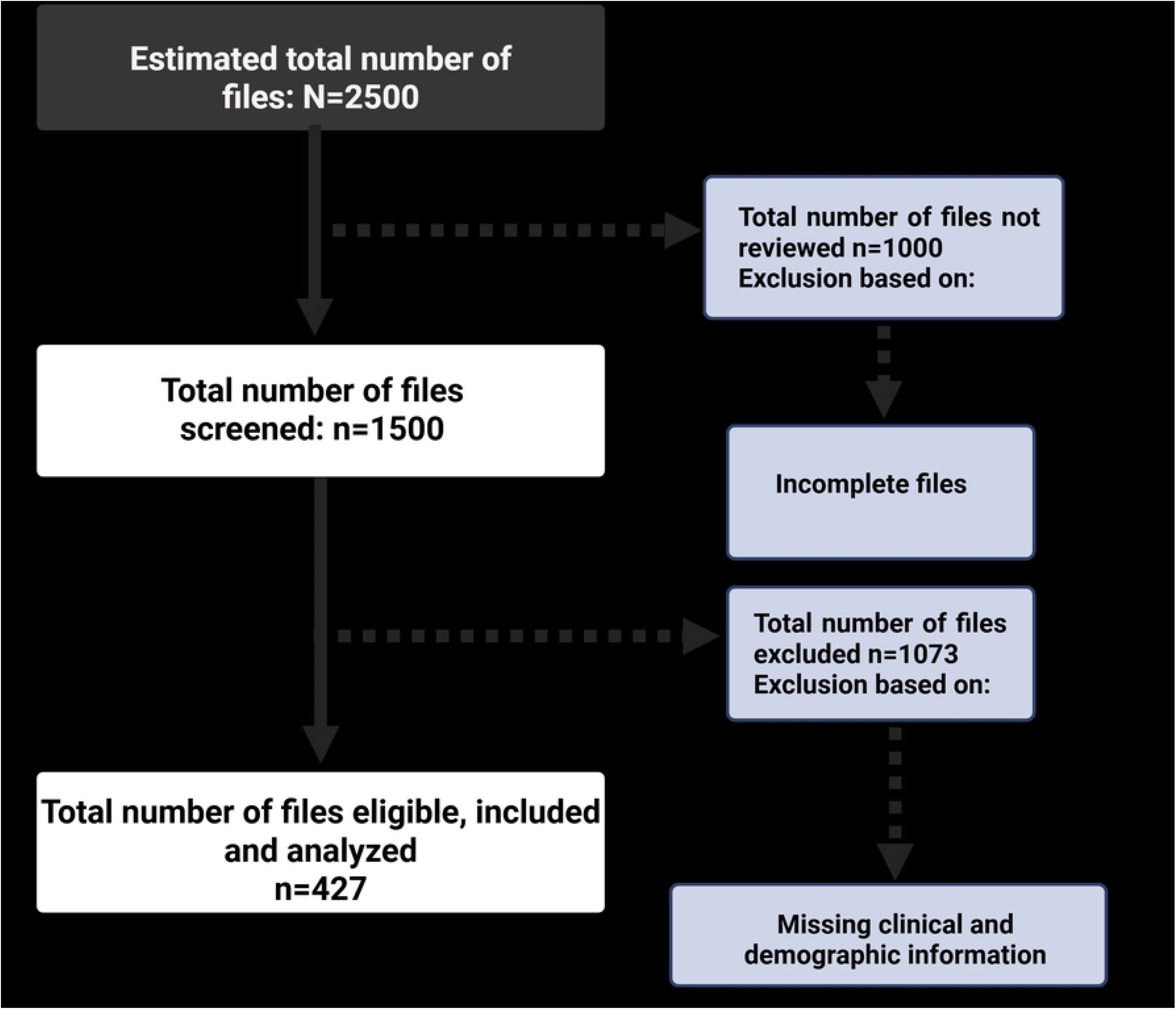
Eligibility Flow Diagram.

The median age of participants was 21 years (IQR: 18–23), with opioid users slightly older (21 [19–23] vs. 21 [18–23], p < 0.0001) **Table 1**. Most participants were male (77.5%), though sex did not significantly differ between opioid users and non-users (p = 0.361). The majority resided in urban areas (57.2%), but residence was not significantly associated with opioid use (p = 0.184). Employment status and marital status also showed no significant differences, though education level approached significance (p = 0.075), with a higher proportion of opioid users in the tertiary education group (33.3%) compared to those with no formal schooling (0%). Opioid use was strongly associated with substance abuse and psychiatric factors. Individuals with alcohol abuse had significantly higher opioid use (15.5% vs. 10.3%, p = 0.0002), while cannabis use showed an even stronger association (76.7% of users vs. 7.8% non-users, p < 0.0001). Psychosis was less common among opioid users (12.1% vs. 19.7%).

Table 2 shows a logistic regression of the relationship between opioid use and other variables. In univariable analysis, alcohol abuse (OR = 2.69, 95% CI: 1.5 - 4.60; p = 0.0003), cannabis use (OR = 3.58, 95% CI: 1.98-6.45; p < 0.0001), and psychosis (OR = 0.56, 95% CI: 0.33-0.95; p = 0.034) were significantly associated with the outcome. After adjustment, systolic blood pressure (AOR = 0.80, 95% CI: 0.65-0.99; p = 0.040) and diastolic blood pressure (AOR = 1.37, 95% CI: 1.03–1.81; p = 0.027) emerged as significant predictors, with alcohol abuse retaining its strong association (AOR = 3.76, 95% CI: 1.47–95874.35; p = 0.036).

**Table 1.** General and clinical characteristics of factors associated with opioid use.

|  |  |  |  |
| --- | --- | --- | --- |
|  | **Opioid** |  |  |
| **Variable** | **Yes =**<br>**66(15.5%)** | **No= 361**<br>**(84,5%)** | **P. value** |
| **Age, Years** | 21 (19, 23) | 21 (18,23) | **<0.0001** |
| **Birth Number** | 3 (2,3) | 2 (1,4) | **<0.0001** |
| **Sex** |  |  |  |
| *Male* | 54 (16.7) | 270 (83.3) | 0.361 |
| *Female* | 12 (12.8) | 82 (87.2) |  |
| **Residence** |  |  |  |
| *Urban* | 39 (10.2) | 179 (47) | 0.184 |
| *Rural* | 21 (5.51) | 142 (37.3) |  |
| **Employed** |  |  |  |
| *Employed* | 5 (20) | 20 (80) | 0.682 |
| *Unemployed* | 36 (16.7) | 179 (83.3) |  |
| **Education Level** |  |  |  |
| *No Formal Schooling* | 0 (0) | 11 (100) | 0.075 |
| *Primary* | 7 (13) | 47 (87) |  |
| *Secondary* | 32 (20.5) | 124 (79.5) |  |
| *Tertiary* | 7 (33.3) | 14 (66.7) |  |
| **Married** |  |  |  |
| *Yes* | 0 (0) | 10 (100) | 0.110 |
| *No* | 167 (79.5) | 43 (20.5) |  |
| **Number Of Children** |  |  |  |
| *0* | 63 (15.7) | 342 (84.4) | 0.631 |
| *1* | 1 (7.7) | 12 (92.3) |  |
| *2* | 2 (40) | 3 (60) |  |
| *3* | 0 (0) | 1 (100) |  |
| *4* | 0 (0) | 1 (100) |  |
| *5* | 0 (0) | 1 (0) |  |
| **Developmental Miles Stones** |  |  |  |
| *Delayed* | 0 (0) | 16 (100) | **<0.0001** |
| *Appropriate* | 235 (100) | 0 (0) |  |
| **Criminal History** |  |  |  |
| *Yes* | 13 (33.3) | 26 (66.67) | **0.0144** |
| *No* | 34 (16.5) | 172 (83.5) |  |
| **Insight Of Mental Illness** |  |  |  |
| *Poor* | 163 (88.1) | 22 (11.9) | **0.025** |
| *Good* | 33 (20.8) | 126 (79.3) |  |
| **Sbp, mmHg** | 116 (108, 127) | 120 (111, 135) | **<0.0001** |
| **Dbp mmHg** | 75 (65, 80) | 75 (67, 88) | **<0.0001** |
| **Alcohol Abuse** |  |  |  |
| *Yes* | 39 (15.5) | 126 (76.4) | **0.0002** |
| *No* | 27 (10.3) | 235 (89.7) |  |
| **Cannabis** |  |  |  |
| *Yes* | 161 (76.7) | 49 (23.3) | **<0.0001** |
| *No* | 17 (7.8) | 200 (92.2) |  |
| **Duration Of Substance Abuse** | 36 (24, 60) | 48 (36, 72) | **<0.0001** |
| **Psychosis** |  |  |  |
| *Yes* | 29 (12.1) | 210 (87.9) | **0.032** |
| *No* | 37 (19.7) | 151 (80.3) |  |
| **Depression** |  |  |  |
| *Yes* | 7 (11.5) | 54 (88.5) | 0.352 |
| *No* | 59 (16.1) | 307 (83.9) |  |
| **Anxiety** |  |  |  |
| *Yes* | 2 (15.4) | 11 (84.6) | 0.994 |
| *No* | 64 (15.5) | 350 (84.5) |  |
| Abbreviations: SBP; systolic blood pressure, DBP; Diastolic blood pressure |  |  |  |

**Table 2.**
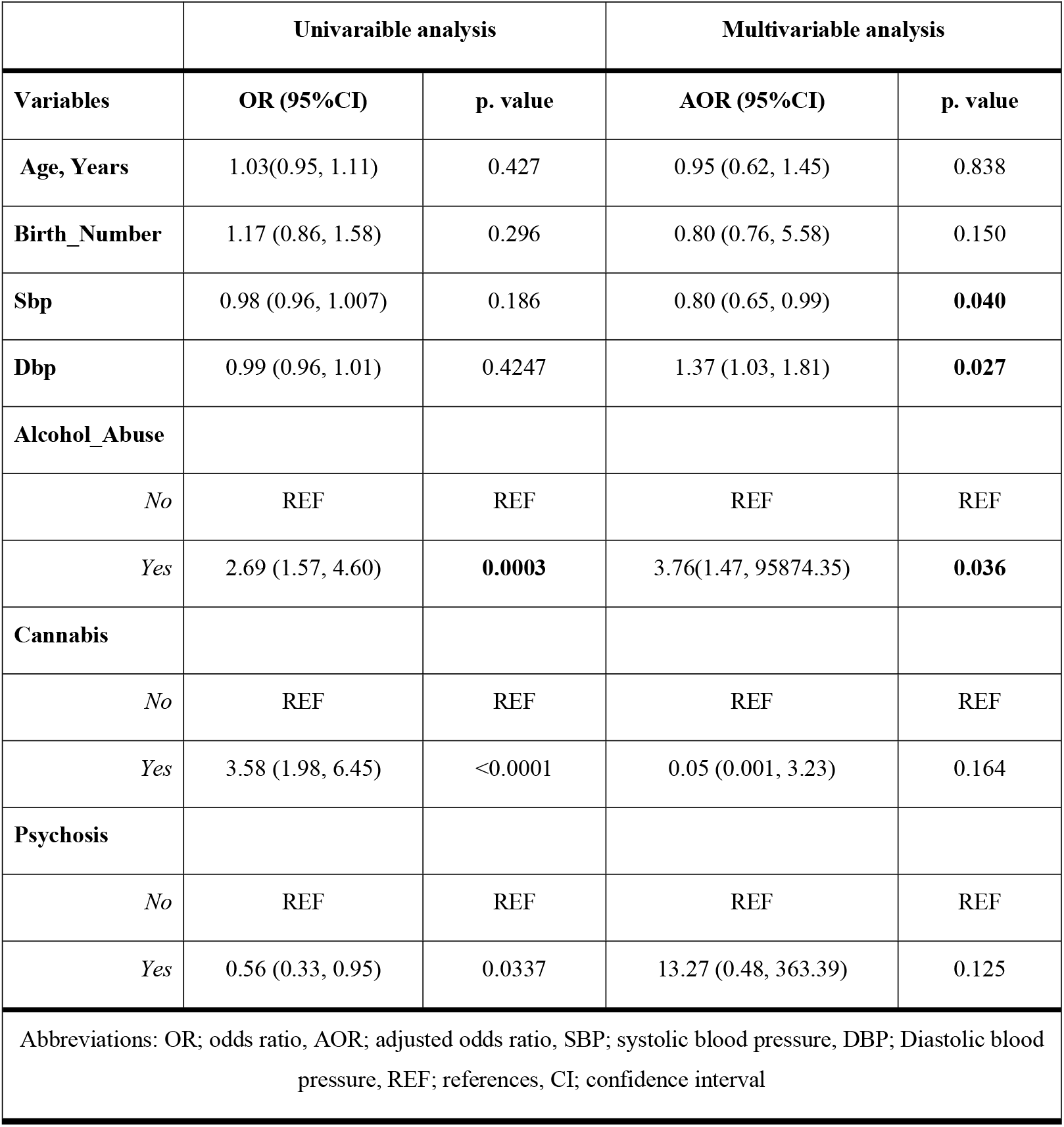
Logistic Regression.

|  | **Table 2. Logistic Regression** |  |  |  |
| --- | --- | --- | --- | --- |
|  | **Univariable analysis** |  | **Multivariable analysis** |  |
| **Variables** | **OR (95%CI)** | **p. value** | **AOR (95%CI)** | **p. value** |
| **Age, Years** | 1.03(0.95, 1.11) | 0.427 | 0.95 (0.62, 1.45) | 0.838 |
| **Birth_Number** | 1.17 (0.86, 1.58) | 0.296 | 0.80 (0.76, 5.58) | 0.150 |
| **Sbp** | 0.98 (0.96, 1.007) | 0.186 | 0.80 (0.65, 0.99) | **0.040** |
| **Dbp** | 0.99 (0.96, 1.01) | 0.4247 | 1.37 (1.03, 1.81) | **0.027** |
| **Alcohol_Abuse** |  |  |  |  |
| *No* | REF | REF | REF | REF |
| *Yes* | 2.69 (1.57, 4.60) | **0.0003** | 3.76(1.47, 95874.35) | **0.036** |
| **Cannabis** |  |  |  |  |
| *No* | REF | REF | REF | REF |
| *Yes* | 3.58 (1.98, 6.45) | <0.0001 | 0.05 (0.001, 3.23) | 0.164 |
| **Psychosis** |  |  |  |  |
| *No* | REF | REF | REF | REF |
| *Yes* | 0.56 (0.33, 0.95) | 0.0337 | 13.27 (0.48, 363.39) | 0.125 |
| Abbreviations: OR; odds ratio, AOR; adjusted odds ratio, SBP; systolic blood pressure, DBP; Diastolic blood pressure, REF; references, CI; confidence interval |  |  |  |  |

## Discussion

Our study aimed at establishing substance use and clinical correlates of opioid dependence among adolescents visiting LUTH. In our study one of the variables that was strongly associated with opioid use was alcohol abuse. The strong independent association of alcohol abuse with the outcome corroborates prior research demonstrating its role in exacerbating chronic health conditions through pathways such as metabolic disruption, immune dysregulation, and poor adherence to medical care [29]. Alcohol abuse, a prominent risk factor identified in this analysis, aligns with evidence from Zambia by matenga et al, where community-based studies have highlighted its role in driving chronic disease burdens and opioid use particularly in urban settings [30]. Local research underscores alcohol’s societal and health impacts, exacerbated by limited access to addiction treatment services and cultural norms surrounding consumption[30,31]. Regionally, similar patterns emerge across sub-Saharan Africa, where alcohol misuse intersects with socioeconomic disparities and healthcare inequities, amplifying its adverse effects [32]. For instance, Rehm et al. (2017) highlighted alcohol’s dose-dependent relationship with multi-organ toxicity in a landmark Global Burden of Disease study, further validating its systemic health impacts[33]. Globally, large-scale epidemiological studies reinforce alcohol’s systemic harms, though challenges in estimating precise effects in smaller or heterogeneous cohorts such as those in low-resource settings emphasize the need for context-specific methodological adaptations [34].

Other variables that were worth mentioning as they proved to show relationship with opioid use were systolic and diastolic blood pressure. The observed relationships between systolic and diastolic blood pressure and the studied outcome reflect a complex interplay of hemodynamic, clinical, and contextual factors, echoing findings from diverse epidemiological and clinical studies. The inverse association of systolic blood pressure (SBP) with the outcome, while seemingly counterintuitive, aligns with paradoxical patterns documented in populations with advanced cardiovascular disease or comorbidities [35]. For instance, studies in aging cohorts have proposed that lower SBP in adjusted models may signal underlying pathologies such as reduced cardiac output in heart failure or compensatory mechanisms in chronic kidney disease, rather than a true protective effect [36]. This phenomenon has been reported in sub-Saharan African settings, where undiagnosed comorbidities like HIV or tuberculosis may confound traditional hypertension risk profiles, as noted in Ugandan and South African research[37].

While the current analysis did not explicitly measure opioid use, existing literature highlights its relevance to DBP dynamics and cardiovascular risk. A Zambian study by Lorenz et al. (2022) reported elevated DBP among individuals using tramadol, a commonly misused synthetic opioid, hypothesizing that its noradrenergic effects may increase peripheral resistance [38]. Similarly, Akande-Sholabi et al. (2019) documented higher DBP in Nigerian cohorts using codeine-containing cough syrups, underscoring the need for region-specific investigations into opioid [39]. These studies align with the current findings on DBP’s role as a risk factor, suggesting that substance-specific mechanisms (e.g., opioid-induced vasoconstriction or autonomic dysregulation) may amplify cardiovascular risk in vulnerable populations.

### Strengths

This study offers significant contributions to the understanding of opioid dependence in a critically understudied population adolescents and young adults in Zambia. By examining sociodemographic, clinical, and physiological correlates within a low-resource setting, the research addresses a pressing gap in global substance use literature. The integration of regional evidence, such as Zambian studies on tramadol misuse and Nigerian data linking codeine syrup to elevated diastolic blood pressure (DBP), enhances the relevance of findings to sub-Saharan African contexts. The use of multivariable logistic regression to adjust for confounders strengthens the robustness of key associations, particularly the strong link between alcohol abuse and opioid use (AOR = 3.76, p = 0.036), which aligns with global patterns of polysubstance use. Additionally, the identification of divergent systolic (SBP) and diastolic (DBP) blood pressure profiles provides novel insights into hemodynamic dynamics in opioid users, a poorly explored area in low- and middle-income countries (LMICs). These findings underscore the value of contextually grounded research to inform targeted public health strategies in regions with limited addiction treatment infrastructure.

## Limitations

Several methodological constraints temper the interpretation of results. The extreme width of the confidence interval for alcohol abuse (95% CI: 1.47–95,874.35) reflects substantial uncertainty in the effect estimate, likely due to sparse data or model overfitting, a common challenge in studies of rare outcomes or heterogeneous populations. This imprecision limits the reliability of quantifying alcohol’s risk contribution. The paradoxical inverse association between systolic blood pressure (SBP) and opioid use raises concerns about residual confounding. For instance, antihypertensive medication use, prevalent in populations with undiagnosed hypertension, or comorbidities like HIV-associated cardiomyopathy common in sub-Saharan Africa may artificially lower SBP independently of opioid use. Furthermore, the cross-sectional design precludes causal inferences, leaving unresolved whether cannabis use predisposes individuals to opioid dependence or vice versa. Reliance on hospital records introduces selection bias, as the sample over represents severe cases (e.g., individuals with psychosis or criminal histories) and excludes community-based populations. Missing data (1,073 excluded records) may further skew results if incompleteness correlates with unmeasured risk factors. Finally, the lack of opioid-specific metrics (e.g., type, dosage, or duration of use) hinders mechanistic insights into how different opioids modulate physiological outcomes. These limitations highlight the need for longitudinal studies with larger cohorts, biomarker integration, and community-based sampling to clarify causal pathways and improve generalizability in future research.

## Conclusion

This study provides critical insights into the complex interplay of opioid dependence, substance use, and cardiovascular health among adolescents and young adults in Zambia, a population often overlooked in global addiction research. The robust association between alcohol abuse and opioid use reinforces the need for integrated substance use interventions in low-resource settings, particularly given the regional rise in polysubstance use and limited treatment infrastructure. The paradoxical inverse relationship between systolic blood pressure (SBP) and opioid use, alongside the positive association with diastolic blood pressure (DBP), underscores the nuanced hemodynamic impacts of opioid exposure, potentially mediated by autonomic dysregulation, undiagnosed comorbidities (e.g., HIV-related cardiomyopathy), or antihypertensive medication effects. These findings align with emerging evidence from sub-Saharan Africa, where tramadol and codeine misuse have been linked to elevated DBP and microvascular dysfunction, highlighting region-specific pathways of cardiovascular risk.

While the study advances contextual understanding, methodological limitations including wide confidence intervals for key predictors (e.g., alcohol abuse), cross-sectional design, and hospital-based sampling bias caution against over interpretation. The extreme imprecision in effect estimates underscores the challenges of studying rare outcomes in small cohorts and calls for larger, longitudinal studies with stratified analyses. Future research should prioritize community-based sampling, biomarker integration (e.g., autonomic function testing), and granular opioid exposure metrics (e.g., type, dosage) to clarify causal pathways. Regionally, partnerships with harm reduction programs and task-shifted hypertension screening could synergize substance use and cardiovascular care, addressing Zambia’s dual burden of addiction and non-communicable diseases. Ultimately, this work highlights the urgency of contextually tailored public health strategies that bridge addiction medicine and cardiometabolic care in sub-Saharan Africa. By grounding global frameworks in local realities, policymakers and clinicians can better address the intertwined epidemics of opioid dependence and hypertension, advancing equitable health outcomes for vulnerable youth populations.

## Data Availability

All relevant data are within the manuscript and its Supporting Information files.

## Availability of data and materials

The raw data underlying the results presented in the study have been uploaded as supporting information.

## Author’s contributions

HH, DNM and SKM conceived the Study. SKM and HH oversaw Data acquisition. SKM and HH supervised data acquisition. SKM and HH conducted the formal analysis. HH,DNM,SMB, KRK, PM,NNN,EY,MN,MM,JK,CS,EL,CM, JPP and SKM wrote the original draft. All authors contributed to the article edits and approved the final manuscript

## Supplementary files

S1. Strobe checklist

S2. Data

